# A Scoping Review of Community-Based Geriatric Health Assessment and Screening Tools used in South Asia

**DOI:** 10.1101/2021.02.19.21252051

**Authors:** Sucharita Panigrahi, Trilochan Bhoi, Sanghamitra Pati, Jaya Singh Kshatri

## Abstract

**Background:** Home-based comprehensive assessment and integrated care of the older people could be a key to relieve the pressure on the already overburdened health system. This review summarises evidence on validated community-based geriatric health assessment tools in South Asia.

**Methods:** Guided by Arksey and O’Malley’s five-stage scoping review framework, a total of 46 studies were included in the scoping review after searching from electronic databases and reference lists using predefined eligibility criteria. Data were extracted on main characteristics of included studies, identified instruments and their psychometric properties of the tools. This review was reported in accordance with PRISMA-ScR guidelines.

**Results:** Among the 46 included studies, 10 reported on instruments for medical assessment, 12 on tools for psychological conditions, 13 on tools for functional issues, 2 on social wellbeing, and 9 on tools with multiple domains of health. Most studies included participants from both gender and different social classes. Majority used measurements that were both self-reported or measured by the investigator. whereas only two instruments were designed to be used by clinicians. In the existing geriatric health assessment tools, environmental domain was neglected completely, and not a single tool considered in this review covered all 5 domains which influence regular life of elderly.

**Conclusion:** There are no validated tools available that can be used for comprehensive geriatric assessment in South Asia. There is a need to develop and validate culturally sensitive tools that can be used for assessing all the geriatric health domains.

## Introduction

An increasingly older population is contributing to a majority of disease burden and subsequently revamping healthcare expenditures globally[1] [2][3]. This burden is more acute in low-income settings where the need for healthcare in older age groups is greater and coverage of health and social security schemes is inadequate[4]. Projections have shown that an overwhelming proportion of global disease burden will be from age-related disorders in the foreseeable future[5].

In this situation, Comprehensive geriatric assessment (CGA) is recognized as a key intervention to address the health needs of older adults, effectively evaluate comorbid conditions and functional limitation[6][7]. Even though, there is no standardized format for carrying out CGA, using multiple tools to capture comprehensive health status, a consensus on the broad domains that need to be measured[8][6]. Most of the tools available focus on a particular domain of CGA and a majority of these have been validated for use in high-income countries(HIC), where they have been developed[9][10][11]. Concerns on the utility and accuracy of these tools in Low and Middle-income countries (LMIC)remain [12]. Unlike clinical evaluation, CGA involves complex constructs that are influenced strongly by the socio-cultural milieu of the end-users and target population, which are widely diversified across regions, let alone the world. Therefore, tools need contextual adaptation and validation for optimum utility in the targeted settings.

While there still are linguistic and regional differences among regions in South Asia, historically, the subcontinent has had a common socio-cultural thread running through it as well as a common administrative architecture, making generalizability of health interventions easier. This scoping review aims to summarize studies that describe validated tools for assessing geriatric health in community settings in South Asia. We believe there is no single tool validated for CGA and there is no structured evidence synthesis conducted to map empirical evidence for tools specifically for this population. So, this review will provide evidence to researchers and practitioners on available validated tools, their psychometric properties, and validity in order to enable them to make an informed choice on which tools to include in community-based CGA. Findings of this review could also aid in identifying scope for updating of existing tools for the South Asia population as well as identify crucial gaps in the domain of CGA where validated tools are missing.

## Methods

This scoping review was reported in accordance with the Preferred Reporting Items for Systematic Reviews and Meta-Analyses statement for reporting systematic reviews-extension for scoping reviews[13] presented in Appendix-1 and was prospectively registered with Open Science Framework(***10*.*17605/OSF*.*IO/TFR3H***).

We followed the Arksey and O’Malley framework [14] for conducting scoping reviews which uses the following steps –

Step1: Identification of Research Question

Step2: Identification of relevant studies

Step3: Study selection

Step4: Data charting

Step5: Data collation, summarize, and reporting the result,

These steps were further refined by Levac et al.[15].

### Identification of Research Question

The research question addressed by this review was developed based on PCC-Population=Geriatric, Concept= Community based health assessment and screening tools, and Context =South Asia.

**R.Q: What are the validated community-based geriatric health assessment and screening tools available in South Asia?**

### Identification of relevant studies

We conducted a systematic search of the following electronic databases: MEDLINE, Embase and PsycINFO. We searched articles published in English only without any date restrictions. The pilot searches were carried out with variations of the words “Community-based”, “elderly”, “Health assessment*” “screening”, “Tool*”, along with countries and states of south Asia that appeared in Title/ Abstracts. The detailed search string is presented in Appendix 2. Additional relevant papers were identified through reference mining and Google Scholar search using similar keywords.

### Study selection

After an extensive search, following de-duplication of electronic articles, all eligible articles were screened by two independent review authors (SP1 & TB) based on their Title/Abstract. Discrepancies were discussed and resolved. In case of disagreement, a third author (JSK) made the decision. Full texts were retrieved and reviewed for eligibility. We included studies conducted in any of the South Asian countries that evaluated geriatric health assessment tools based on community setting, or any of the subdomains or reported its development process or validation. Institutional tools which can be implemented in the community were also included. We excluded manuscripts that were in form of comments, editorials, letters, Conference or congress papers, abstracts, and reviews. Full-text review followed the same method as Title/abstract screening in case of disagreement. In this study, we have not considered methodological rigor of the included studies.

### Charting the data

Through iterative process, a predefined data extraction sheet was developed to capture all relevant aspects of the included studies. The study characteristics like country of origin, objectives, sample characteristics, setting, sample size, and sampling method were extracted. Detailed description of the health assessment tools and the procedure of its development, composition, validity, reliability, and feasibility of the instruments were extracted from the relevant paper. After completion, the charted tables were examined further within the reviewers to ensure accuracy and consistency. The detailed descriptions of the studies are given in appendix-3.

### Collating, Summarising, and reporting the results

A narrative summary of the results was presented. Tables were used to present specific details of the tools and development process. The results section first described the characteristics of the studies, characteristics of the tool, and its psychometric properties. The charted tools were categorized into five broad domains and further subdomains of CGA as follows[16][17].

a. Medical Assessment
b. Functional Assessment
c. Social Assessment
d. Environment Assessment
e. Multiple

Similarly, we also summarized the most reported outcome measures of validity and reliability such as sensitivity, specificity, positive predictive value (PPV), negative predictive value (NPV), and receiver operating characteristics area under the curve (ROC-AUC). We have grouped all the available tools with psychometric properties in 3 categories to make our results standardized.

1. High sensitivity/specificity: When the tool reported value is ranging from 67%-100%.
2. Moderate Sensitivity/Specificity: When the tool reported value is ranging from 34%-66%
3. Poor Sensitivity/ Specificity: When the tool reported value is ranging from 0%-33%

## Results

We identified 607 records from electronic databases and other sources after de-duplication. 116 were identified for full-text screening and 46 studies were included in the final analysis. The PRISMA flow diagram for the study selection is provided in Figure 1.

**Figure 1:**
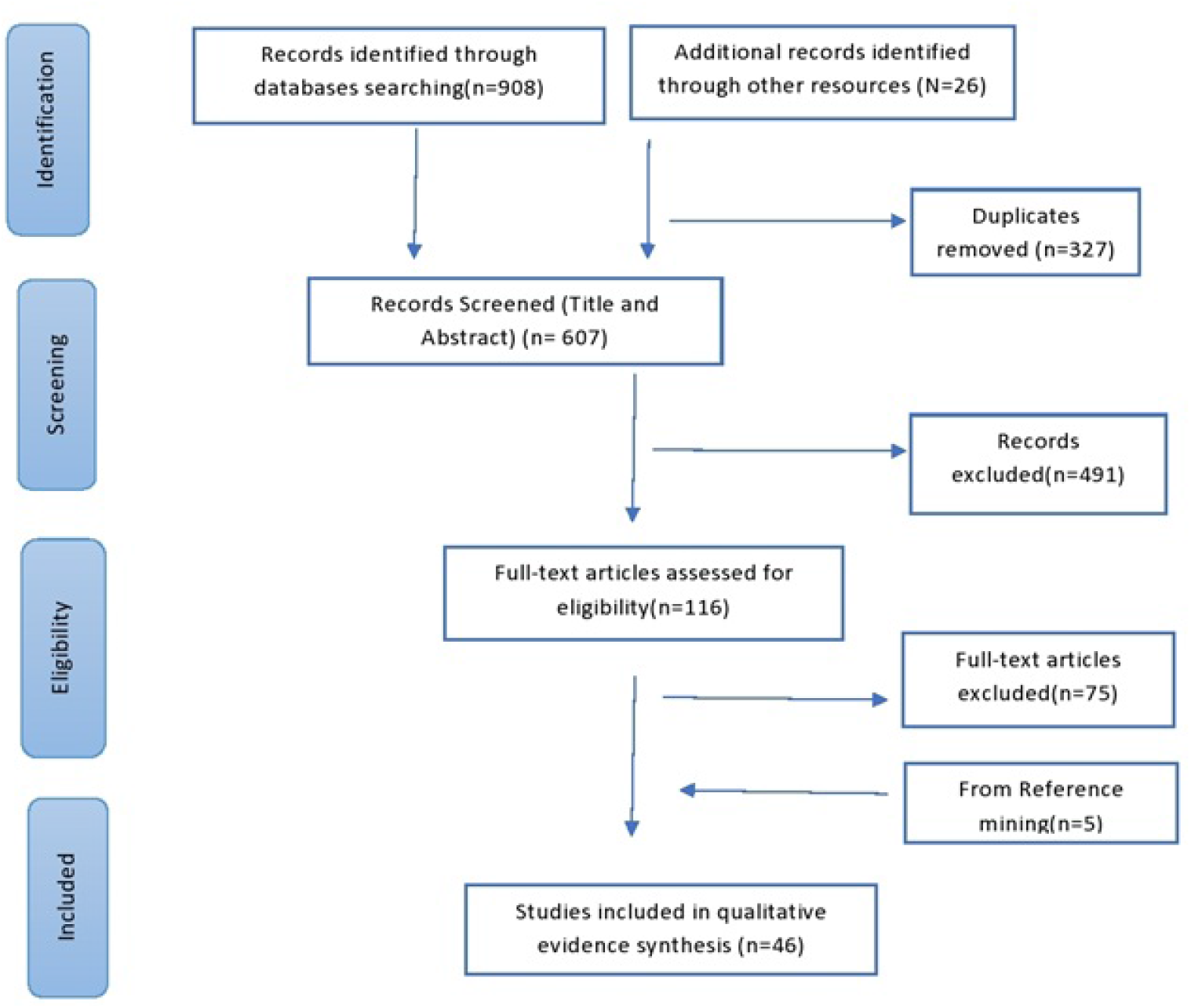
PRISMA flowchart of the Geriatric Scoping Review

All studies were cross-sectional and aimed to either develop, validate or test tools for assessment of different health parameters. A considerable number were part of larger studies, but a few were multinational in scope, so we are presenting the components that concerned the tool development/validation in South Asia only. All studies included participants from all genders, different age groups and social classes, except Rathnayake-2020, who included only postmenopausal women[18]. The study characteristics are summarized in Table-1 below.

**Table-1:**
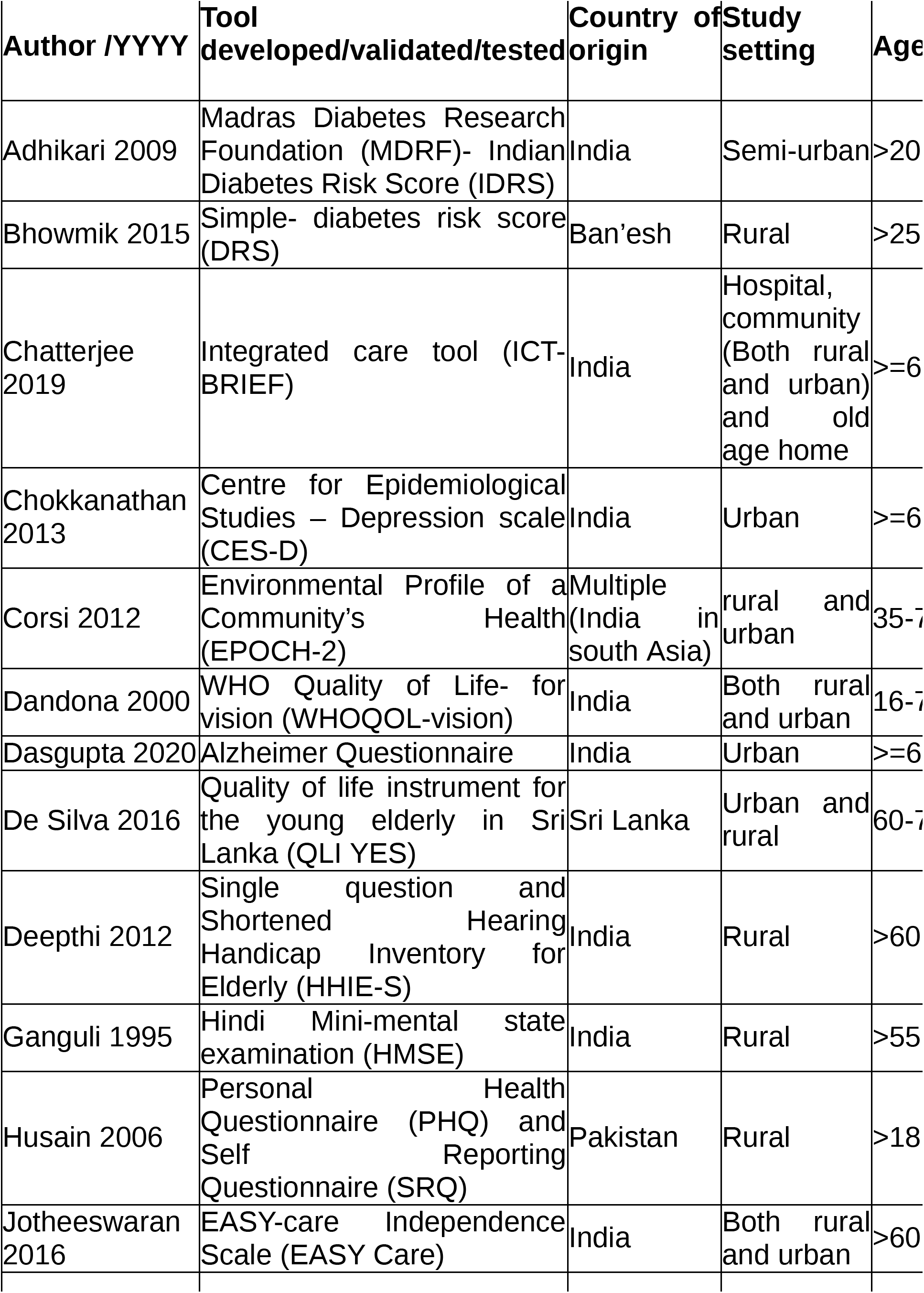
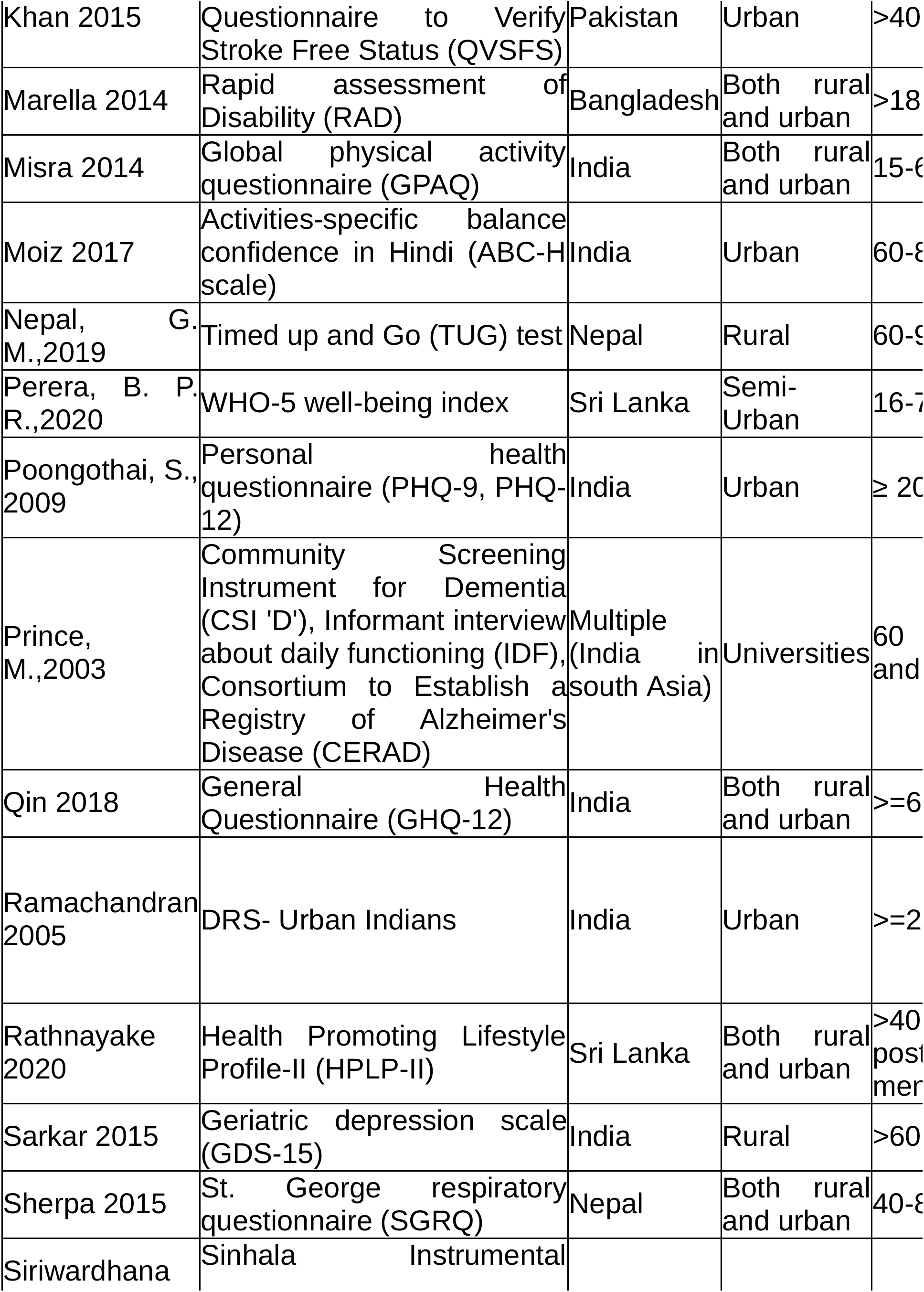
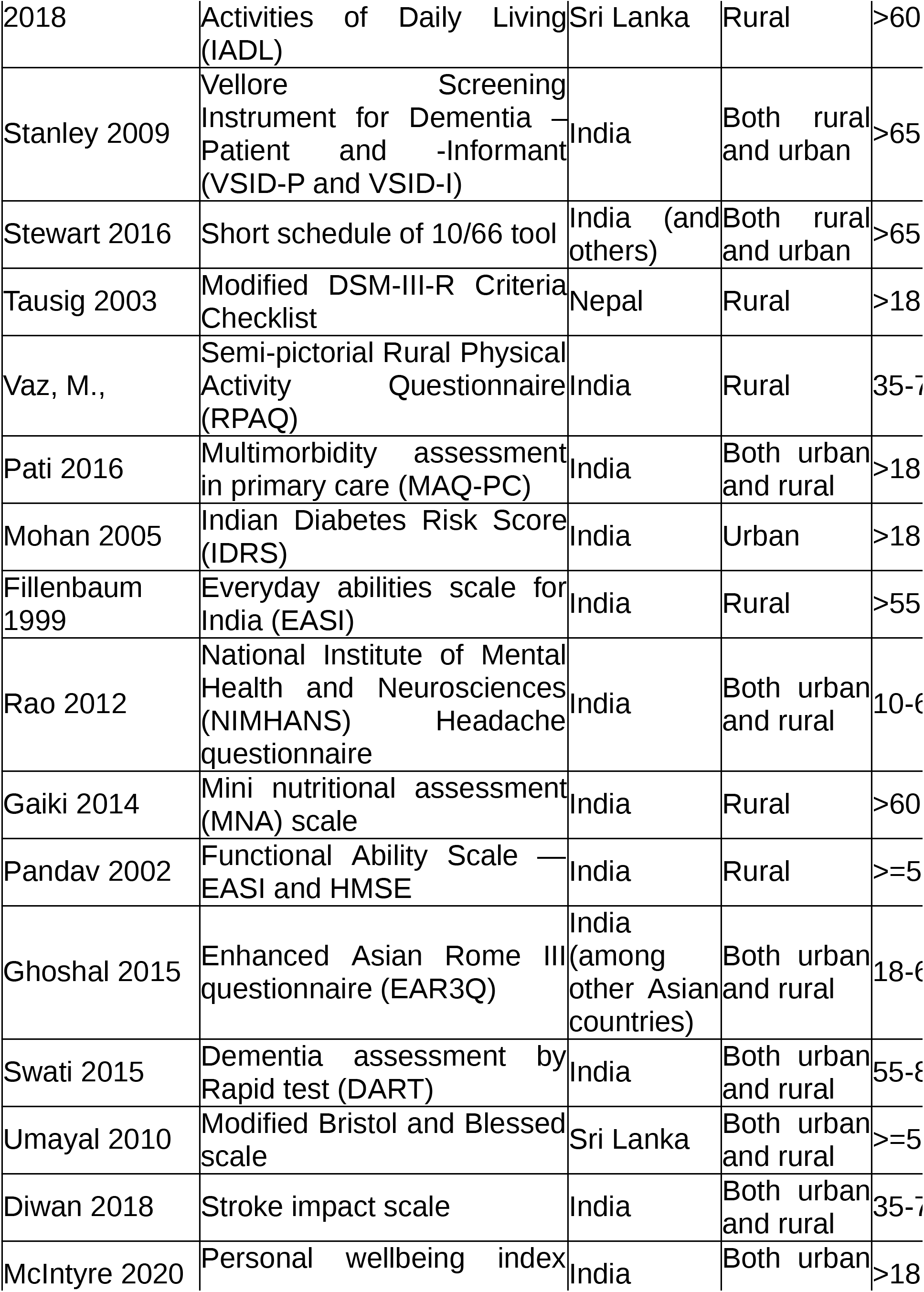
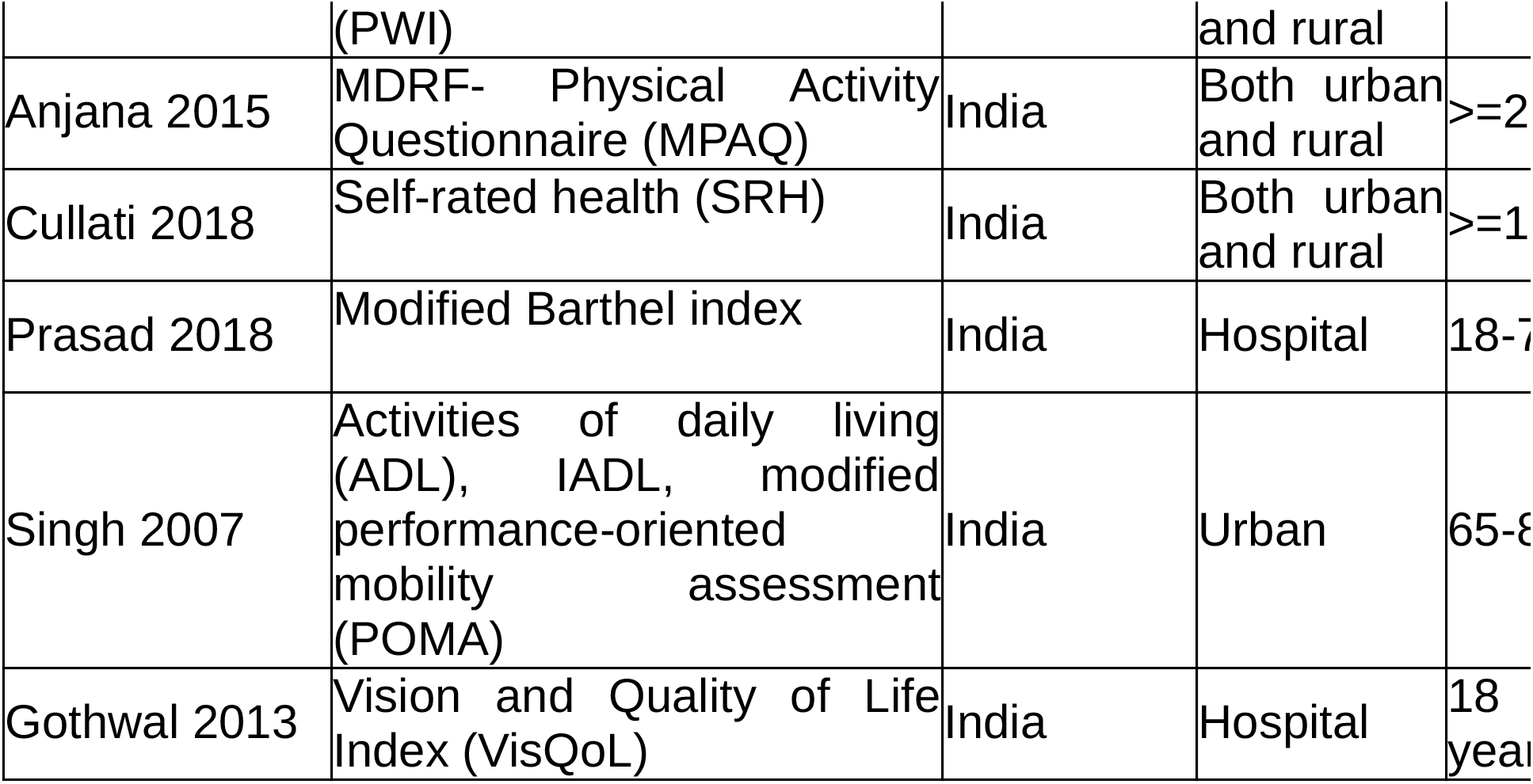
Characteristic of Included studies.

All tools were either self-administered or administered through an investigator or health worker, except two tools, modified DSM-III checklist and EASY-care [20], which were designed to be used by clinicians. The characteristics of the tools are provided in Table-2 below.

**Table-2.**
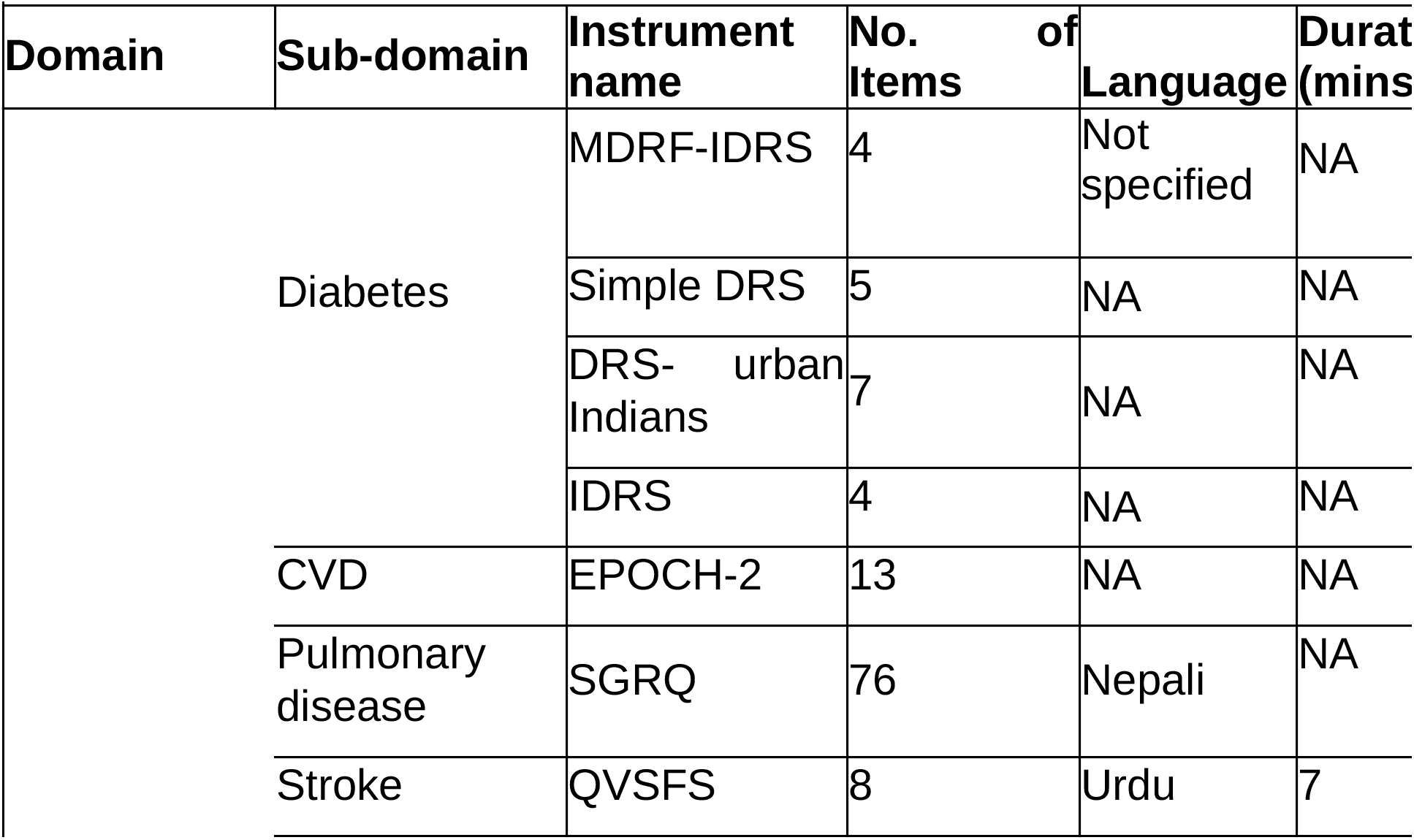
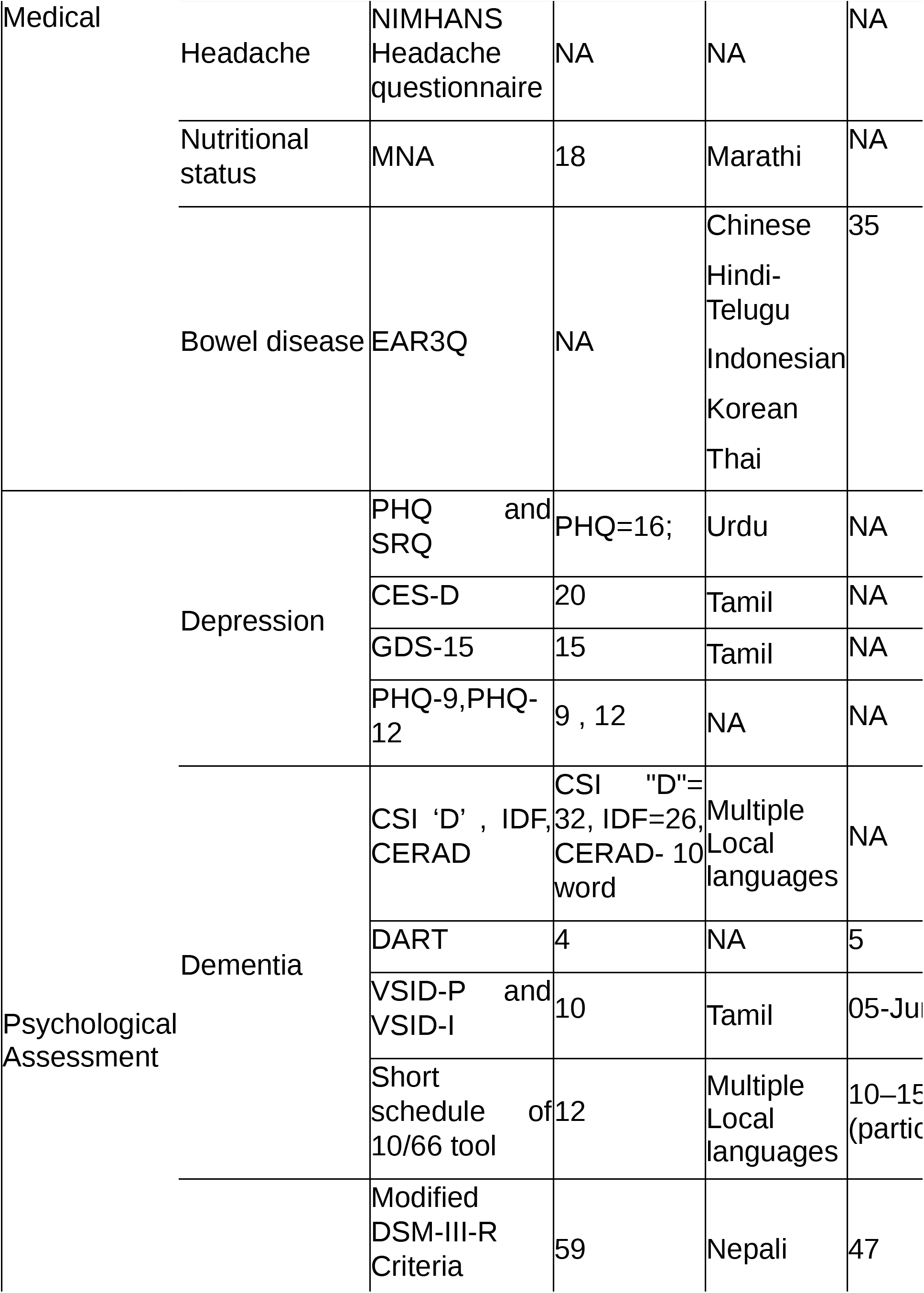
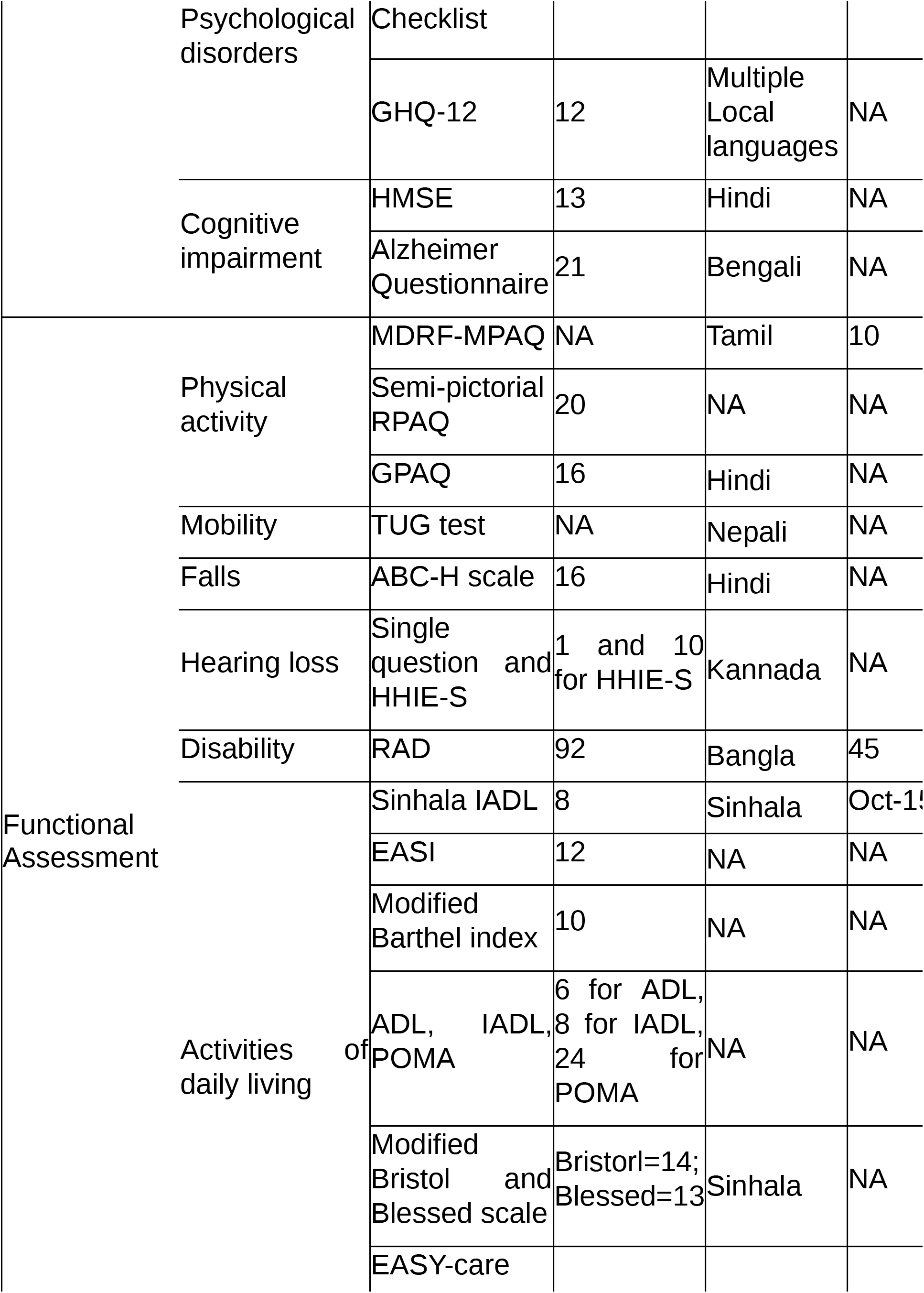
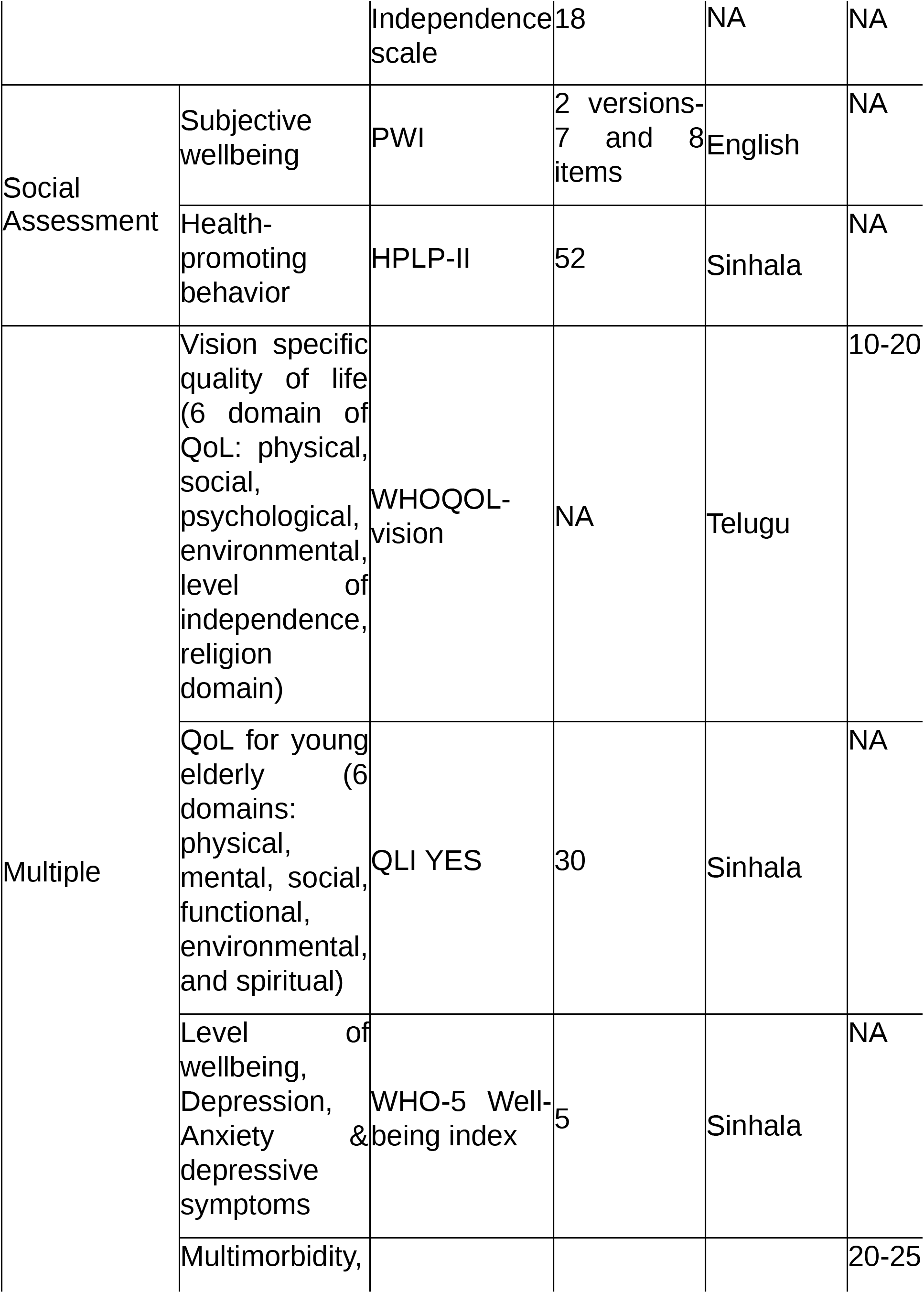
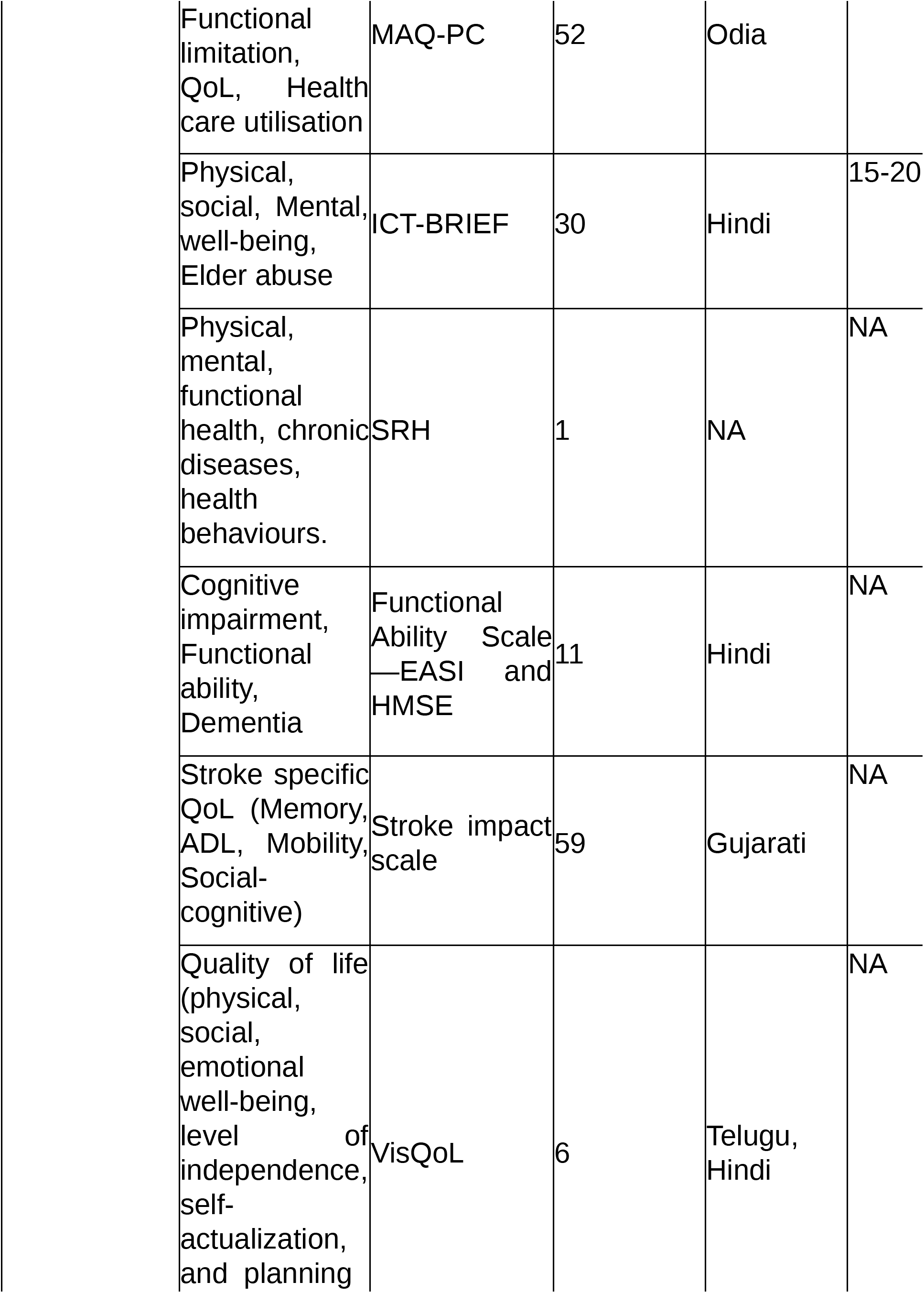
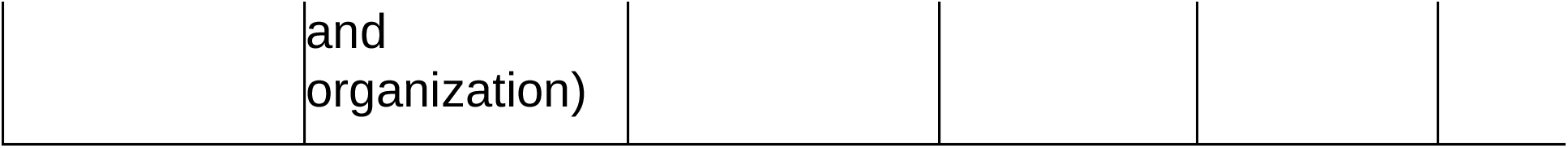
Characteristics of the Tools.

### Measures of tools

Majority of the tools were compared with a defined “Gold standard” to evaluate the sensitivity, specificity, predictive values, and cut-off optimization, which was reported in some form for 24 tools and summarized in table-3 below. The other common measures of tool design and validation reported were factorial analysis by Bartlett’s test of sphericity, Kaiser-Meyer-Olkin (KMO) measure of sampling adequacy, exploratory and confirmatory factor analysis (EFA/CFA), content, convergent, divergent, criterion and construct validity, inter-item and spearman’s correlation, reliability by tests of internal consistency such as Cronbach’s α and test-retest reliability by intraclass correlation coefficients (ICCs), inter-rater reliability by Cohen’s Kappa.

The available tools were categorized into five major domains: -

#### A. Medical Assessment tools

The included studies on 10 tools related to medical assessment in older adults. Four tools - MDRF-IDRS[21], Simple DRS[22], DRS-urban Indians[23], IDRS[24] were for risk scoring of diabetes which had showed moderate sensitivity and specificity. The QVSFS[25]was intended to verify the stroke-free status, NIMHANS Headache questionnaire[26] for headache and EAR3Q[27] for bowel disease, all these tools had high sensitivity and NPV. The EPOCH-2[28] and MNA[29] were assessed cardiovascular and nutritional status respectively and reported moderate reliability for both. SGRQ[30] used to predict COPD and had high negative correlation between SGRQ scores and lung capacity.

#### B. Psychological assessment tools

There were four studies each that assessed tools related to depression (PHQ/SRQ[31],CES-D[32], GDS-15[33], PHQ-9-12[34]) and dementia (CSI ‘D’/IDF/CERAD[35], DART[36], VSID-P/VSID-I[37], Short schedule of 10/66 tool[38]). All of these reported moderates to high sensitivity and specificity as shown in table 3.

**Table 3.**
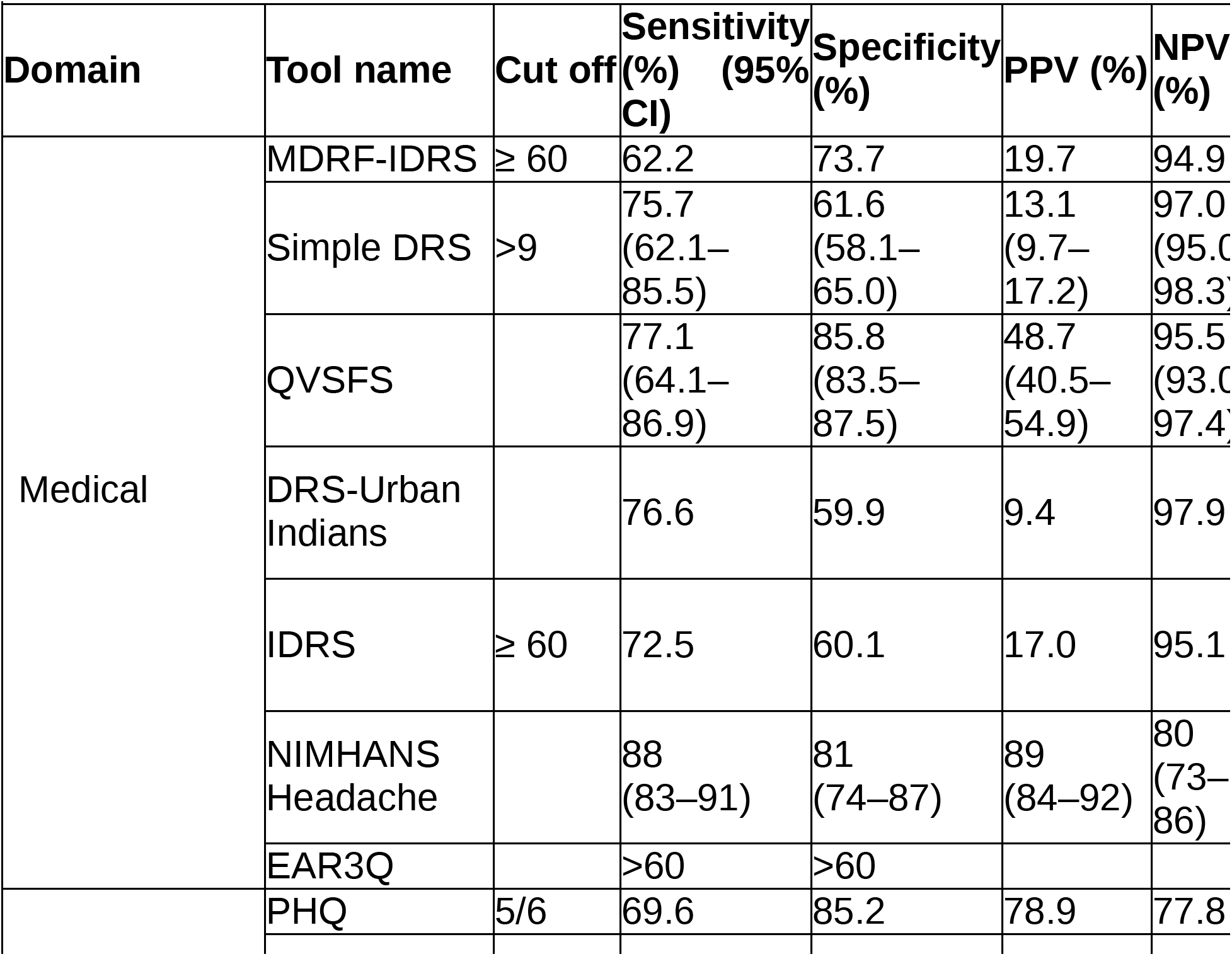
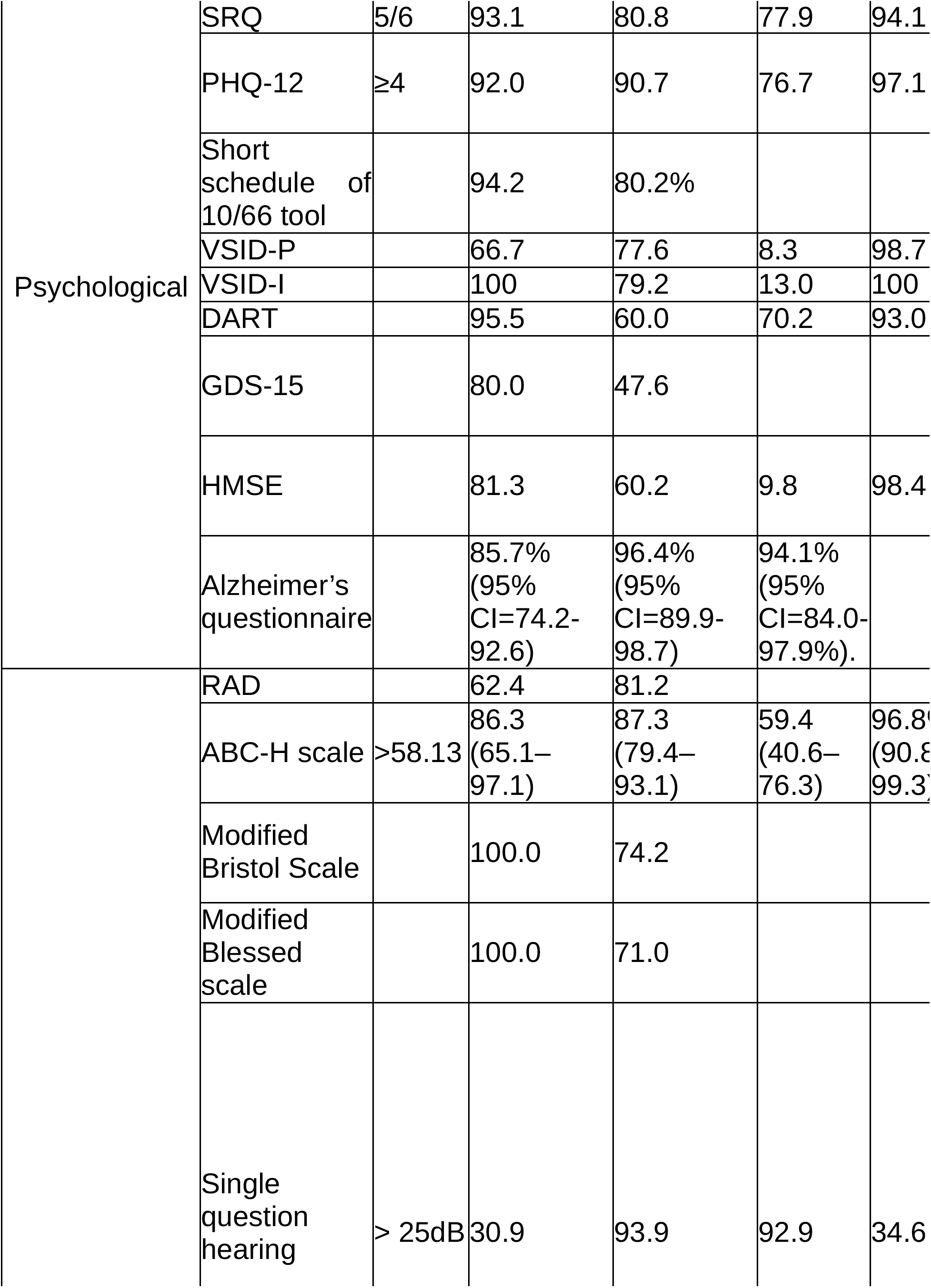
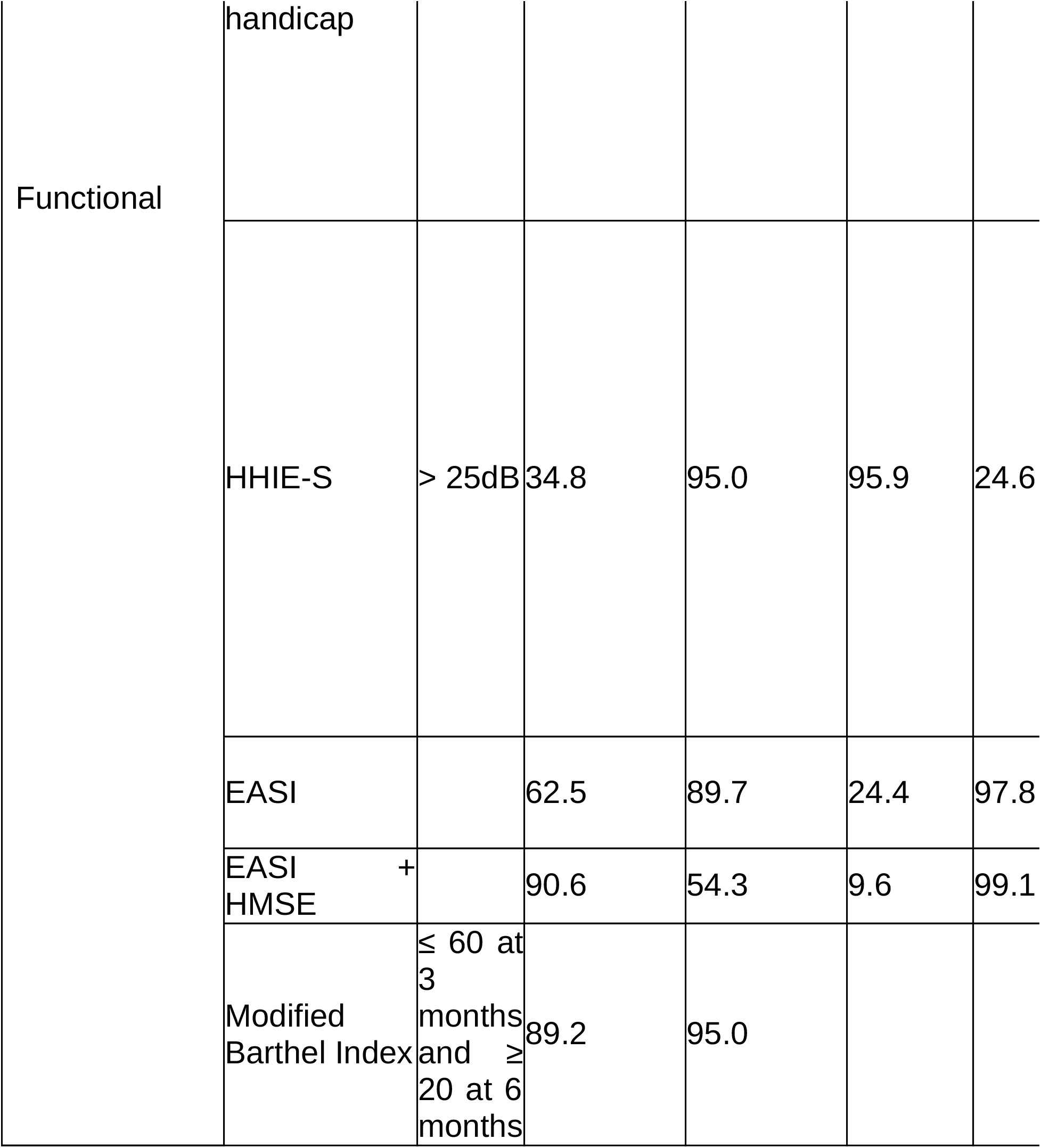
psychometric properties of the Tools.

The DSM-III[19] checklist had moderate construct validity, variable reliability with poor alpha values for schizophrenia and mania, along with high alpha values for depression and anxiety. Whereas, the GHQ-12[39] tool showed moderate reliability. In case of HMSE[40] had moderate sensitivity and specificity, high NPV but poor PPV. However, Alzheimer’s questionnaire[41] showed high sensitivity, specificity and high PPV.

#### C. Functional assessment tools

There are five tools (Sinhala version of ADL[42], Modified Barthel index[43], EASI[44], EASY Care[20], modified Bristol and Blessed scales[45]) were validated for ADL assessment. Among them, Sinhala version of ADL[42] and EASY Care[20] independence scale had high reliability, Whereas Modified Barthel index[43], substantial-high sensitivity and specificity, EASI[44] tool had shown moderate sensitivity and high specificity, with a poor PPV and the modified Bristol and Blessed scales[45] had very high sensitivity and moderate specificity.

RAD assesses disability with moderate sensitivity and high specificity. ABCH scale to assess falls had high sensitivity and specificity with a high PPV. Single question hearing handicap and HHIES both used for hearing loss, had poor sensitivity and high specificity with poor NPV.

#### D. Social assessment tools

Subjective wellbeing as assessed by PWI[46] and health-promoting behavior assessed by HPLP-II[18]. Both tools had high reliability.

#### E. Multiple domain assessment tools

A total of nine tools assessed more than one domain and all had some form of a quality-of-life assessment included in them. The WHOQOL vision[47], VisQoL[48], The QLI-YES[49] included 6 different domains of quality of life and reported high to moderate reliability.

The WHO-5 well-being index[50] is used to assess and compare depression, anxiety & depressive symptoms. The translated version demonstrated good content and face validity with high reliability.

The MAQ-PC[51] tool assessed multimorbidity, functional limitation, depression, health-related quality of life and health care utilization. It had an overall moderate to high reliability and validity. ICT-BRIEF tool[52] and SRH[53] tools to assess different domains of quality of life had high reliability and t moderate validity respectively

Stroke impact scale[54] assessed the stroke-specific QoL and reported moderate positive correlations between constructs of memory, communication, ADL, mobility, and hand function. It showed weak positive correlation with participation and physical domains.

The psychometric properties of the tools described above are summarized in Table-3 and also available in supplementary file (Appendix-3).

## Discussion

This systematic scoping review aimed to describe available tools to assess geriatric health in community settings in South Asia. We found that there is a paucity of tools that can be used to evaluate elderly health comprehensively. Majority of tools available evaluate specific health outcomes, rather than broader conceptualizations of complete wellbeing and hence are not suitable for use in assessment of multiple domains of health. A large majority(29 instruments) were adopted from pre-existing instruments, mostly developed for the western population. This indicates insufficient research and uptake of health measurements and scales particularly for the South Asian population. While two studies were from before 2000, many studies (32) were published in the past decade. Research aimed at tool development and validation in South Asia is probably a relatively recent phenomenon.

A combination of steady declining birth rate and increasing life expectancy has been seen in South Asia that has resulted in an increasing aging population, along with an increasing burden of morbidity. This amplifies the need for CGA and the tools necessary for its implementation., This would play an important role to decide individual needs and thus provide person-centered care. This review also reported medical and functional health assessment seems to be the major domains of CGA represented by maximum number of tools rather than the whole spectrum of health. While assessing geriatric QoL, environment plays an essential role, but tools to assess this domain are the least frequently found. Studies conducted on elderly population in China have shown a clear association between environmental factors and survival, where air pollution was shown to have a significant association with elderly health[55]. Similarly, tools assessing financial burden of illness, access to health care or social health were not found.

The priority in the region has traditionally been on addressing infectious diseases, maternal and child health challenges. Elderly health has therefore remained a neglected area of policy, practice and subsequently research in the region. Although few targeted health programs such as the “National Programme for Health Care of the Elderly (NPHCE)” in India [56] have been initiated in recent years, their impact is yet to be evaluated. These programs envision geriatric assessment and screening in their activities but the uptake has been slow due to unavailability of tools for the same that can be used by the community health workers.

Most of the programs in developing countries have attached less importance to home-based elderly care because of the emphasis given to institutional, mostly medical care. This finding is supported by our review where only one tool MAQ-PC[51] included most of the domains (Medical, Psychological, Functional) of CGA but that too considering the institutional-based care needs of the participants.

Health systems need to appreciate and be prepared for the shift from communicable to non-communicable diseases in the coming years. An increasing focus on healthcare for the elderly in the coming decades is inevitable. Tools for the health assessment and screening of this group need to be contextually developed and validated in multiple settings to realize the goals of the national programs.

### Limitation of the review

Only peer-reviewed articles published in English language were selected, which may introduce selection bias, however, it is also important to note that the primary language for scientific communication in the whole of South Asia is English. We have not evaluated the quality of the studies included. We have limited our studies to South Asia, this could increase the chance of missing out the studies conducted in other LMICs. However, the compliance of the review with PRISMA guidelines with robust and systematic methodology strengthens the confidence in findings.

## Conclusion

This review indicated a significant gap in tools available for CGA in the South Asian population that can be used in community settings. No tool combining all aspects of CGA has been reported from the region. Acknowledging the significance of context-specific robust health assessment tools, there is a need for culturally sensitive and practical tools that can be used to assess geriatric health.

## Data Availability

Data will be provided on request.

## Authors’ Contributions

SP1 & TB were equally contributed to the development of search strategy, screening, hand searching, charting, and analysis of the data. SP1 drafted the article. JSK was responsible for conceptualization, manuscript review, and editing.SP2 was responsible for manuscript review and editing. All the authors have read and agreed to the published version of manuscript.

## Funding

This review received no external funding.

## Conflict of Interest

The authors declare no conflict of interest

## Notes

### Competing Interest Statement

The authors have declared no competing interest.

### Funding Statement

Nil

### Author Declarations

Not applicable. This scoping review registered in Open Science Framework (OSF) prior to beginning of the study.

