## Appendix-2 for "A Scoping Review of Community-Based Geriatric Health Assessment and Screening Tools used in South Asia"

Search Strategy

Search terms used in databases MEDLINE, Embase and PsycINFO

1.Community-Based Participatory Research/ or Community participation/ or (Community-Based Participatory Research* or Community-based research* or Community-based or communit* or Community-dwelling* or population-based or Community participation* or Community-directed).ti,ab,kw.

and

2.Aged/ or (Aged or older people or elderly people or Older adult* or Elderly or Aging or Ageing or Geriatric* or Senior* or older person*).ti,ab,kw.

and

3.Health impact assessment/ or (Health impact assessment* or Health assessment* or Assessment*).ti,ab,kw. Or diagnosis/ or "diagnostic techniques and procedures"/ or (Diagnos* or Examination*).ti,ab,kw. Or Geriatric assessment/ or Geriatric assessment*.ti,ab,kw. Or Mass Screening/ or (Mass Screening* or Screening*).ti,ab,kw. Or (Instrument* or Tool* or Scale* or Index* or Questionnair* or Evaluation*).ti,ab,kw.

and

4.Validation study/ or (Validation* or Validity or Reliability or Development*).ti,ab,kw.

and

5.Afghanistan/ or Bangladesh/ or Bhutan/ or India/ or Nepal/ or Pakistan/ or Sri Lanka/ or (Afghanistan or Bangladesh or Bhutan or India or Nepal or Pakistan or Sri Lanka or South Asia or Maldives).ti,ab,kw.
